# Mobile Monitoring of Mood (MoMo-Mood): a Multimodal Digital Phenotyping Study with Major Depressive Patients and Healthy Controls

**DOI:** 10.1101/2024.06.27.24309600

**Authors:** Talayeh Aledavood, Nguyen Luong, Ilya Baryshnikov, Richard Darst, Roope Heikkilä, Joel Holmen, Arsi Ikäheimonen, Annasofia Martikkala, Kirsi Riihimäki, Outi Saleva, Ana Maria Triana, Erkki Isometsä

**Author notes:** Corresponding Author: Talayeh Aledavood, PhD, Computer Science Aalto University, 02150 Espoo Finland.

## Abstract

**Background:** Mood disorders (MD) are among the most common mental health conditions worldwide, significantly contributing to mortality, morbidity, and disability rates. In today’s society, individuals generate digital traces through their interactions with wearable and consumer-grade personal digital devices. These traces can be collected, processed, and analyzed, offering a unique opportunity to quantify and monitor individuals with mental disorders in their natural living environments. Various forms of digital traces from the use of smartphones and other personal digital devices hold the potential to reveal new behavioral markers associated with depressive symptoms in patients with mood disorders.

**Objective:** We conducted an observational longitudinal study, MoMo-Mood Pilot, involving a cohort of patients with major depressive disorder (MDD) and a healthy control group without any diagnosed mental disorder to confirm the feasibility of our digital phenotyping study design. Upon confirming feasibility, we conducted a year-long, more extensive study, MoMo-Mood, with a similar design. This study comprised (1) three subcohorts of patients with a major depressive episode (MDE), including those with either MDD, bipolar disorder (BD), or concurrent borderline personality disorder (BPD), as well as (2) a healthy control group without any diagnosed mental disorder. We investigated whether differences in behavioral patterns, as quantified by passively collected digital trace data, could be observed at the group level, i.e., patients vs. healthy controls. We studied the volume and temporal patterns of smartphone screen and application usage, communication, sleep, mobility, and physical activity. Additionally, we examined which of the passively quantified behavioral features and sociodemographic factors are associated with the presence of depression in study participants at different points in time.

**Methods:** MoMo-Mood Pilot and MoMo-Mood recruited 201 participants. The pilot study enrolled 14 patients with MDD and 23 healthy controls. The participants completed a 2-phase study: the first two weeks (the *active phase*) involved collecting data from bed sensors, actigraphy, and smartphone data passively collected through the Niima platform. Additionally, participants had to actively engage with the study by answering five sets of questions daily, prompted on their smartphones throughout the day. In the second phase (the *passive phase*), which lasted up to 1 year, only passive smartphone data and bi-weekly Patient Health Questionnaire (PHQ-9) assessments were collected. The MoMo-Mood study, which had a similar setting to the pilot study, enrolled 164 participants: 133 were patients diagnosed with current MDE through structured interviews (85 with MDD, 27 with BPD, and 21 with BD), and 31 were healthy controls. Survival analysis was performed to compare the adherence of healthy controls and the patient subcohorts. Statistical tests were performed to compare behavioral patterns, and a generalized linear mixed model was used to assess the association between different factors and the presence of depression.

**Results:** The study design was proven feasible upon completing the pilot study. Survival analysis using the log-rank test showed no statistically significant difference in participants’ adherence between subcohorts. The overall communication volume was similar in both groups, but the weekly temporal patterns revealed distinct preferences as the controls typically made or received calls in the afternoon. In terms of location patterns, significant differences emerged: weekday location variance showed lower values for patients (Control = -10.04 ± 2.73, Patient = -11.91 ± 2.50, MWU test *P*-value = .004) and normalized entropy of location was also lower among patients (Control = 2.10 ± 1.38, Patient = 1.57 ± 1.10, MWU test *P*-value = .05). However, no differences were observed in weekend location patterns. The temporal communication patterns for controls were found to be more diverse than that of patients (MWU test, p<0.001). In contrast, patients’ smartphone usage temporal patterns were more varied than the controls. Investigating mobile-derived behaviors and their relationship with the concurrent presence of depression, we observed that the duration of outgoing calls over the past two weeks was negatively correlated with the presence of depression (beta = -7.93, 95% CI: -13.17 to- -2.69, *P*-value = .003). Conversely, longer durations of incoming calls (beta = 5.33, 95% CI: -0.08 to 10.75, p = 0.053) and a larger proportion of time spent at home (beta = 5.16, 95% CI: 0.78 to 9.54, *P*-value = .002) were positively associated with the presence of depression.

**Conclusions:** This work demonstrates the design of a longitudinal digital phenotyping study harnessing data from a cohort of patients with depression. It also shows the important features and data streams for future analyses of behavioral markers of mood disorders. However, among outpatients with mild to moderate depressive disorders, their group-level differences from healthy controls in any of the modalities alone remain overall modest. Therefore, future studies need to be able to combine data from multiple domains and modalities to detect more subtle differences, identify individualized signatures, and combine passive monitoring data with clinical ratings.

## Introduction

Mood disorders, including major depressive disorder (MDD) and bipolar disorder (BD), are substantial contributors to the worldwide disease burden and impairment, bearing significant implications for public health [1,2]. They also have an increased association with premature mortality [3,4]. Mood disorders are usually recurring and often chronic conditions that typically require long-term treatment [5]. Currently, the diagnostic landscape and assessment of symptom severity for mood disorders are primarily based on clinical evaluations, interviews, and questionnaires, predominantly reliant on subjective interpretations and retrospective recollections by patients and only conducted when the patient visits the mental health professionals. This framework is susceptible to memory bias and may not capture the dynamic symptomatology inherent in mood disorders [6,7]. Compounded by the lack of clear biomarkers and a worldwide shortage of adequate mental health resources, the treatment of these conditions has long posed challenges.

In recent years, the ubiquity of digital technologies, particularly wearables and personal digital devices, has facilitated their integration into different fields of medicine. These devices can measure various physiological metrics such as heart rate, heart rate variability, and respiration rate [8]. Moreover, through interactions with these devices, significant amounts of digital traces are generated, which can be collected, analyzed, and transformed into valuable information regarding their users’ behavior and activities. In the case of psychiatric disorders, particularly mood disorders, leveraging digital traces and the associated information holds the potential to introduce greater objectivity in diagnosing and treating these conditions as well as the possibility for continuous monitoring of patients. The emerging field of digital phenotyping harnesses real-time indicators of human behavior to uncover behavioral markers that objectively quantify, monitor, and assess mental health conditions continuously [9–14].

Several digital phenotyping studies have used data collected from personal digital devices to extract behavioral features. These studies typically employ clinically validated scales–such as the Patient Health Questionnaire-9 (PHQ-9) [15]–to measure symptoms of the disorder, using them as a reference point. Most digital phenotyping studies aim to determine which passive behavioral features, gathered as participants interact with their devices, are most indicative of clinical scores. A recent review [16] examined 45 articles and found that most of these studies focus on a single modality. In contrast, a multimodal approach could provide a more accurate understanding of an individual’s life and behaviors. Also, many of the studies involve non-clinical cohorts or individuals who self-identified as depressed through self-administered questionnaires rather than diagnosed patients.

In response to significant challenges in digital phenotyping, we designed and ran the Mobile Monitoring of Mood (MoMo-Mood) study. These challenges include the scarcity of research involving clinically diagnosed patients, the necessity for prolonged observation periods to identify behavioral precursors to recurrent episodes in mood disorders, and the requirement for a multimodal approach to capture behaviors from various data streams. This year-long, multimodal digital phenotyping study focused on different types of patients who were receiving treatment for major depressive episodes, including (unipolar) MDD, BD, or borderline personality disorder (BPD), as well as healthy controls. Before MoMo-Mood, we ran a smaller pilot study (MoMo-Mood Pilot) to test the feasibility of our study design before scaling it to a larger number of participants and patients with less prevalent disorders. In this work, we explore the potential and effectiveness of personal digital devices in capturing clinically significant behavioral indicators from diagnosed patients over one year. The unique combination of patient subcohorts, a wide range of data streams from different devices, and an extended study time allows us to, for the first time, test the feasibility of such a study design at a larger scale and gain insights into what works and what needs to be improved for future studies.

While study adherence has been recognized as a challenge in digital phenotyping studies [17], how different groups of patients comply with long-term passive data collection has yet to be well-studied. Similarly, while past research has suggested that patients with serious mental disorders have similar or even higher rates of smartphone usage compared to others [18,19], the frequency and quantity of smartphone usage in different groups of mood disorders need to be better examined. To this end, we measure adherence based on the time each person has remained in the study (maximum one year). Also, we use smartphone screen activity (on, off, lock, unlock) timestamps as a proxy for general smartphone usage and smartphone applications (apps) usage for specific types of usage.

In addition to that, we hypothesize that patients (compared to healthy controls) exhibit differences when comparing activities and behaviors that are relevant for assessing mood disorder symptoms or associated psychosocial disability. Here, we focus on four domains: social activity, mobility, physical activity, and sleep. Depression is known to be associated with social isolation [20], which can potentially be reflected in a person’s communication pattern. Fatigue and loss of energy are among the diagnostic criteria for depression [21,22], which can be reflected in mobility and physical activity data. Past research has used passively collected location data from smartphones. It has identified variables such as location variance, location entropy, and time spent at home to be associated with symptoms of depression [23–25]. Finally, both increased and decreased sleep are symptoms of depression [26]. In our study, as a proxy of social activity, we use time stamps for incoming and outgoing calls and text messages (SMS) through the phone network provider. Location and mobility are measured through the smartphone’s GPS and WiFi data, and the variables shown to exhibit significant differences between patients and healthy controls in other studies are investigated. We measure physical activity in two ways: first, through actigraphs, and second, through smartphone accelerometers. Sleep timing and duration are measured by actigraphs and bed sensors.

Moreover, to assess the number of different behaviors, we focus on their temporal patterns by exploring activity rhythms and how patients and controls allocate their time to a particular activity throughout the week. The timing of different activities may contain important behavioral markers linked to mood disorders. Previous research in studying temporal rhythms of activity inferred from different personal digital devices has shown that people tend to have persistent activity rhythms over time [27–30]. However, these rhythms and their consistency are linked to personal and sociodemographic characteristics [31,32]. A recent study [33] suggests that daily activity rhythms inferred from smartphone GPS data are associated with anxiety levels in patients with MDD and bipolar disorder. While we know that mood disorders impact daily activity and sleep rhythms, the activity rhythms of patients with mood disorders inferred from passively collected data from their smartphones are not well studied [34]. This work investigates the different activity rhythms inferred from smartphones and evaluates the differences between patients and healthy controls.

We specifically aim to answer the following research questions (RQs):

**RQ1** Does study adherence, i.e., how long the subjects remain in the study, vary between patient subcohorts and healthy controls?

**RQ2** Do the quantity and temporal patterns vary between patients and controls?

**RQ3** Are the differences between patients and healthy controls when comparing social activity, mobility, physical activity, and sleep as quantified from passively collected data?

**RQ4** Are there within-group differences in temporal rhythms of smartphone usage social and physical activity?

**RQ5** Is the presence or absence of depression during follow-up associated with social activity, mobility, physical activity, and sleep?

This study identified significant differences in usage patterns and the predictive value of smartphone-derived features for depressive symptoms among patients and healthy controls. Although daily communication volume and overall smartphone usage did not differ between the groups, their temporal patterns of smartphone use varied. Specifically, controls typically made or received calls in the afternoon, whereas patients tended to do so in the morning. Additionally, controls displayed a greater diversity in the timing of their communication and physical activities. Significant variances were also observed in location patterns during weekdays, particularly regarding location variance and entropy. Furthermore, our analysis confirmed that the duration of incoming and outgoing calls, along with the proportion of time spent at home, are predictors associated with the presence of depressive symptoms.

Our results can help future studies improve their designs, increase adherence, and focus on the most promising digital traces and meaningful features. Ultimately, these findings, combined with insights from future similar studies, may lead to the development of tools for monitoring and in-time intervention for patients with mood disorders.

## Methods

### Setting

We initially ran a smaller-scale pilot study, the MoMo-Mood Pilot, to test our study design. Once the study was deemed feasible, we moved on to a larger-scale study, namely MoMo-Mood. Other than the number of participants, the main difference between the two studies was that for the Pilot study, participants were either MDD patients or healthy controls, as opposed to the main study, where two additional patient subcohorts were recruited. The recruitment of all the cohorts for the main study was performed independently from the pilot study.

Both studies were run in Finland. Patients who had already been diagnosed using structured clinical interviews were recruited on a voluntary basis from the Mood Disorder outpatient treatment facilities of the Helsinki University Hospital Mood Disorder Division, Turku University Central Hospital Department of Psychiatry, and City of Espoo Mental Health Services. The Mini International Neuropsychiatric Interview [35] was used for the diagnosis of MDD and BD, and the Structured Clinical Interview-II [36] was used for BPD. The patients with any psychotic features, concurrent substance use disorder, or imminent risk of suicide were excluded.

The studies had an active phase that lasted two weeks, during which we collected data from actigraphs and bed sensors loaned to study participants, and different types of data were passively collected (passive data) from smartphone sensors through a smartphone application. Passive data is a byproduct of the participants’ use and engagement with personal devices; they do not need to change their behavior or actively engage with the study. During the active phase, we also did ecological momentary assessment (EMA) [37]five times a day. In the second phase of the study, the passive phase, which lasted for up to a year, passive data was collected from smartphones (Table 1.) However, actigraph, bed sensor, and EMA data were no longer collected. Participants got a prompt every two weeks on their smartphones to fill out a PHQ-9 questionnaire. They were also presented with PHQ-9 at baseline. The division of the study into active and passive phases was implemented to minimize the risk of study fatigue and burden on participants.

**Table 1:**
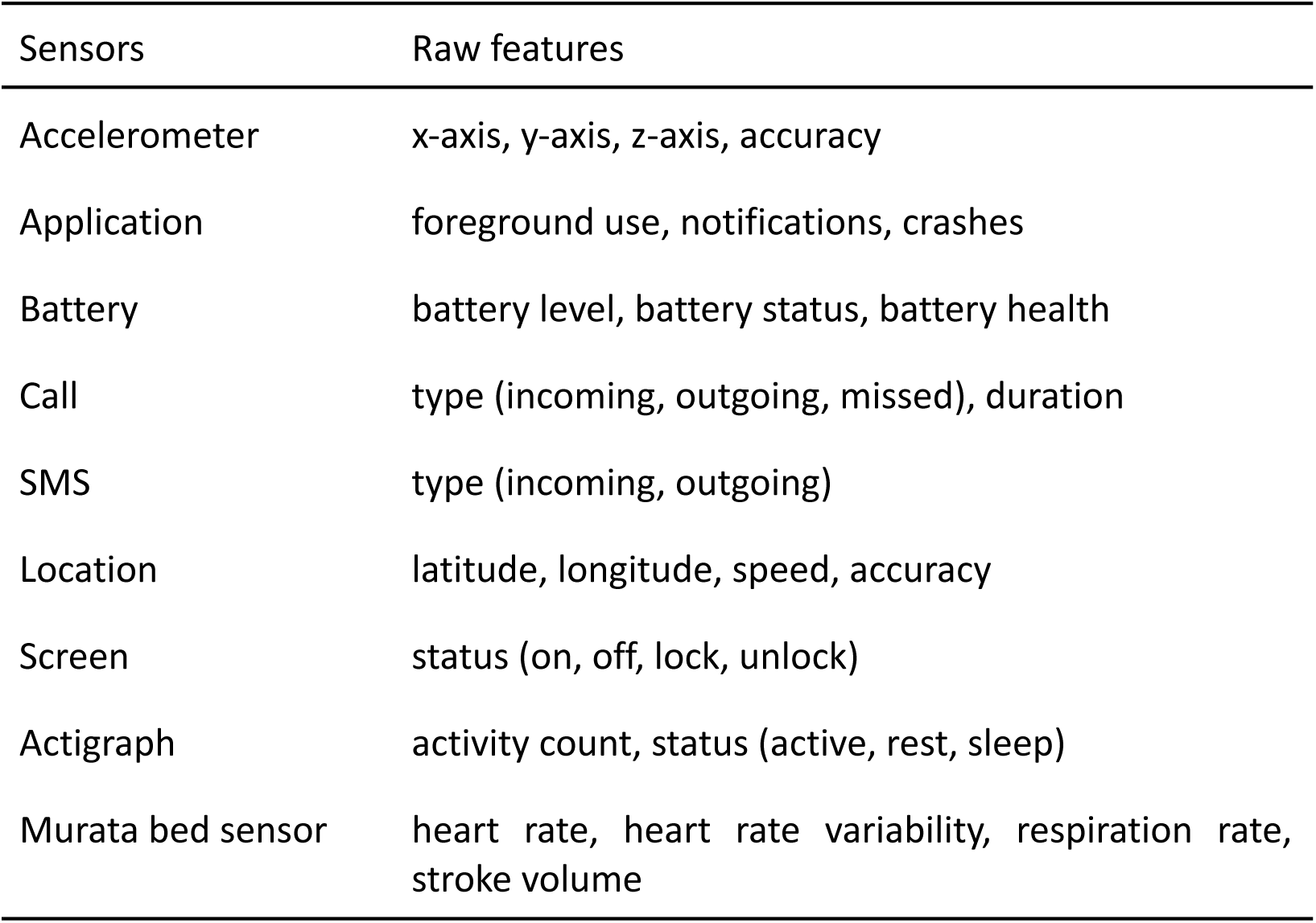
Raw features description.

| Sensors | Raw features |
| --- | --- |
| Accelerometer | x-axis, y-axis, z-axis, accuracy |
| Application | foreground use, notifications, crashes |
| Battery | battery level, battery status, battery health |
| Call | type (incoming, outgoing, missed), duration |
| SMS | type (incoming, outgoing) |
| Location | latitude, longitude, speed, accuracy |
| Screen | status (on, off, lock, unlock) |
| Actigraph | activity count, status (active, rest, sleep) |
| Murata bed sensor | heart rate, heart rate variability, respiration rate, stroke volume |

The actigraphs we used did not need to be recharged for 14 days, allowing us to provide fully charged devices to participants without recharging during the active phase. Additionally, PHQ-9 asks about depression symptoms in the past two weeks, which made the two-week period a natural choice for the length of our active phase. Given the time frames of when people typically change their smartphones or the operating system on the smartphones, we chose the length of one year for the passive phase of the study. The participants could withdraw from the study at any time. Figure 1 demonstrates the study’s timeline.

**Figure 1:**
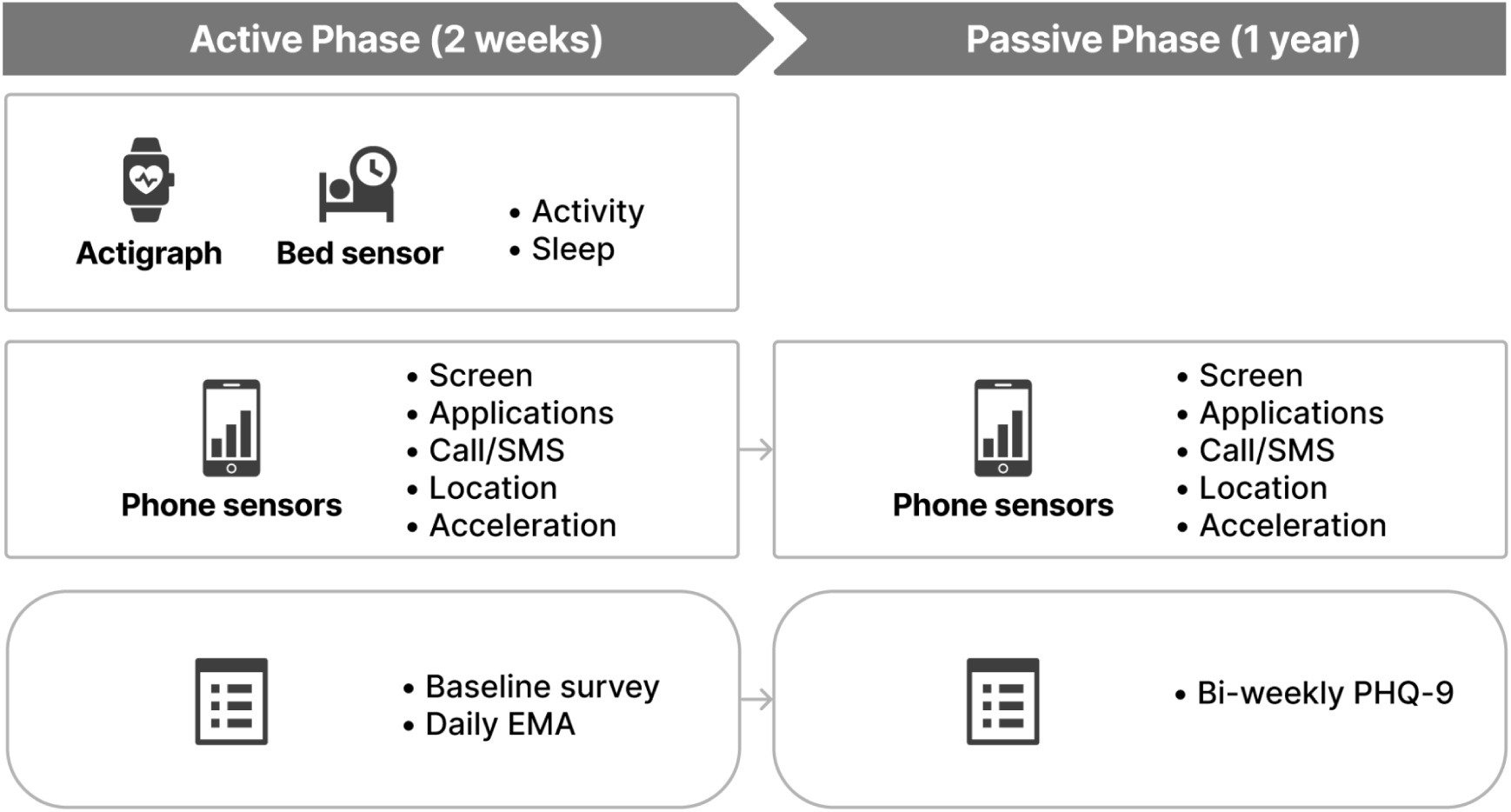
Data collection procedure of the MoMo-Mood study.

### Participants and Recruitment

#### MoMo-Mood Pilot

23 healthy controls and 14 patients with MDD were recruited. One control was excluded from the study for having a baseline PHQ-9 score that fell in the sub-clinical depression range. The healthy controls were 17 females and 5 males, with an average age of 30.5 ± 10.4 years. The patients were 8 females and 5 males. The average age for the patients was 32.4 ± 12.2 years. One patient had not entered their age and sex information in the forms. In the pilot study, for practical reasons, we asked patients to stay in the study for six months and the healthy controls for one year. This work refers to the MoMo-Mood Pilot study as the *Pilot dataset*.

#### MoMo-Mood

In this study, a total of 164 participants from 4 different groups: 1) healthy controls (N=31), 2) patients with MDD (N=85), patients with BPD (N=27), and patients with BD (N=21) were recruited. The participants were invited to stay in the study for a maximum of one year. The length of involvement, thus data provided by each subject varied widely, from days to the entire year. Participants were enrolled continuously, entering and leaving at different times. This work refers to the MoMo-Mood study as the *Main dataset*.

#### Data Collection Platform

The Non-Intrusive Individual Monitoring Architecture (Niima) is a data collection platform designed for MoMo-Mood Pilot and other digital phenotyping studies [38]. This platform allows for the design of studies with various parameters and links data from different sensors. It also provides basic cleaning and preprocessing to standardize data into tabular formats for easy handling and analysis. The Niima platform was used to run both studies.

### Data Sources

#### Smartphone

We used the AWARE phone application [39] to collect data from the smartphone sensors. Our team modified and adapted this open-source framework to cater to the needs of our studies. Data collected from smartphones included communication (call and text) timestamps, an anonymous identifier for contacts, timestamps of smartphone screen events (on, off, lock, unlock), GPS data, application use, and battery status. Each record included the user’s ID, the device’s ID, and a timestamp. The smartphone also served to ask participants several questions multiple times a day during the active phase of the study and to present them with the PHQ-9 questionnaire every two weeks during the passive phase. Table 1 summarizes all the raw features gathered from each sensor.

#### Ecological momentary assessment (EMA)

In clinical studies, EMA measures real-time emotional states [37]. In this method, participants respond to questions about their current state on their smartphones multiple times daily. In our studies, a prompt appeared on each participant’s smartphone once in the morning, once in the evening, and three times at random points in time (within specific ranges) throughout the afternoon. These questionnaires asked participants about their psychological and physical states, physical activity, behavior, and sleep. This paper focuses on the behavioral patterns measured objectively through passive data. Results from the EMA data are reported elsewhere [40].

#### Actigraph

Actigraphs are wrist-worn devices commonly used in clinical studies (including psychiatric studies) to measure activity levels and sleep. We used Philips Actiwatch 2 devices. The data was aggregated at 30-second intervals.

#### Bed Sensor

We employed ballistocardiography-based Murata SCA11H nodes (bed sensors) to measure bed acceleration. Participants were provided with these sensors and a pre-configured WiFi router, enabling data transmission directly to the study server. The sensors locally analyzed data, providing heart rate, heart rate variability, respiration rate, stroke volume, and signal strength at 1 Hz frequency.

### Data Preprocessing

#### Communication and smartphone screen usage

The number of incoming/outgoing calls, SMS, and screen use episodes were extracted hourly. The mobile application did not transfer message content or identities of contacts off of the device. To ensure equal evaluation of event volume and temporal patterns, we retained participants with at least eight weeks of communication and smartphone usage data for further analyses. Records from the first and last day were excluded to avoid incomplete data capture. No additional outlier filtering was performed. To analyze the weekly temporal patterns or "rhythm" of events, we aggregated these events into one-hour bins across eight weeks, resulting in 168 bins for weekly frequency. One event is one instance of a particular data type, such as one call or smartphone screen-on data point. The aggregated values were normalized to sum up to 1, providing a proportionate measure of event distribution over time.

#### Applications

The frequency and duration of smartphone applications (apps) usage were collected based on timestamps when apps were actively used in the foreground. Each app was manually categorized to reflect its function, including: “sports,” “communication,” “leisure,” “social media,” “work,” “news,” “shop,” “transport,” and “utility.” App usage durations larger than 10 hours (132 instances) were removed as they might represent an error in the system rather than actual app usage. The weekly rhythm of app usage was computed using the same approach for communication and screen time.

#### Accelerometer

Accelerometer data was recorded every 30 seconds. The magnitude was computed as the Euclidean norm of the acceleration vectors (x-axis, y-axis, and z-axis) and normalized to center around the zero value. We used the standard deviation of accelerometer data at hourly intervals as a proxy for physical activity, similar to prior work [41,42]. Additionally, physical activity was estimated using actigraph data described below.

#### Sleep and activity detection

Sleep variables were derived from actigraph and bed sensor data. For actigraph, sleep/wake epochs were scored by Oakley’s algorithm [43]. Each epoch was assigned a status indicating whether the user was active, at rest, or asleep. The active hours for each day were calculated based on the total number of epochs labeled as “active.” Bedtime was determined as the first occurrence of a continuous three-minute sleep period.

Similarly, wake time was determined as the first instance of the first three-minute span of activity observed in a day. For bed sensors, each 1-second epoch was given a status indicating whether the bed was occupied or free. The sleep period was counted as the longest duration of which the bed was occupied. For both sensors, records on the first and last nights were excluded, and nights with less than 3 or more than 13 hours of sleep were omitted [44]. The preprocessing step resulted in 862 actigraph days and 841 bed sensor nights.

#### Location and mobility

Several mobility features were extracted from the GPS sensor using the Niimpy behavior data analysis toolbox [45]. The choice of features is motivated by previous studies [23,24]. Preprocessing details and feature descriptions are provided in Appendix 1.

### Statistical Analyses

Descriptive statistics were used to summarize the demographic characteristics of participants. Survival analysis was performed using the Kaplan-Meier curve [46] to assess the likelihood of staying in the study over time for different groups. The log-rank test was used to compare the differences in adherence between groups. This non-parametric test is commonly used in survival analysis in the presence of censored data.

The comparative analysis of the quantity of communication and smartphone usage among different groups was performed using the Mann-Whitney U (MWU) test. Results with a *P*-value smaller than 0.05 were considered significant.

A generalized linear mixed model [47] was used to investigate the association between passive data and the presence of depression for repeated measurements. Participants’ responses were divided into two groups based on PHQ-9 score: no or mild symptoms = PHQ-9 < 10, severe = PHQ-9 >= 10. We built a model for each sensor using the group as response variables and sensor measures as predictors. The mean values of sensor measures in the 14-day window preceding a PHQ-9 measurement were computed. All predictors were min-max normalized.

### Temporal Analysis

#### Weekly activity rhythms

To determine how an individual distributes a specific activity temporally, we build *weekly activity rhythms.* The rhythm was computed as the normalized distribution of the aggregated volume of variables for eight weeks. Intra-difference of rhythm was defined as the cosine distance between the average rhythm of individuals belonging to the same cohort,

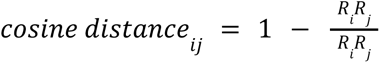

*R_i_*⋅*R_j_* represents the dot product of the activity distributions of individuals *i* and *j*, *||R_i_||* and *||R_j_||* are these distributions’ Euclidean norms (or magnitudes).

Intuitively, individuals with similar rhythms will share a smaller distance. MWU was conducted to compare mean differences in intra-group distances to compare between-group rhythm diversity. Post-hoc correction using the Benjamini-Hochberg procedure [48] was used to control for the False Detection Rate during multiple tests.

### Ethical Considerations

The research protocol for both MoMo-Mood Pilot and MoMo-Mood was approved by the Helsinki and Uusimaa Hospital District’s (HUS) Ethics Committee and was granted research permits by HUS Psychiatry. The approvals covered data streams, data collection platform security, and participant consent information. All data in transit was encrypted, and participant privacy was a key design value. Local IT support and the research ethics committee approved the written data security statement. Participants received detailed information about the study and the data to be collected prior to enrollment. All participants provided written consent. Participants were informed that they could opt out at any point in time. The only compensation they received was four movie tickets each.

### Data Availability

Due to the highly sensitive and private nature of this data, the data collected in these studies cannot be shared with researchers outside of our consortium. Our research permit does not allow the free availability of these data.

## Results

The main goal of the MoMo-Mood Pilot study was to prove the feasibility of our study design. This study had no significant issues, which led us to continue to the main study with minimal changes overall. The results presented here focus on the main study; we compare them with the pilot study where applicable.

We describe the general statistics of participants in both studies and answer the five research questions. The first research question (RQ1) compares study adherence between different subgroups. RQ2 investigates whether there are differences in the smartphone usage patterns of patients and controls to assess the difference in technology usage between groups. RQ3 focuses on symptoms or associated psychosocial disability related to depression. We use passive data as a proxy to measure and quantify social activity (communication), sleep, physical activity, location, and mobility. In RQ4, we investigate the temporal rhythms of phone usage, communication, and physical activity to study whether patients or controls exhibit more homogeneous activity patterns within their group. Lastly, RQ5 uses repeated measurements for the presence of depression and investigates which passive data are associated with the presence of depression.

### Participants

In the main study, a total of 164 participants were recruited. Due to technical challenges or other issues, 13 participants provided no passive data and were excluded from all analyses. Further analyses included 30 control subjects and 121 participants diagnosed with MDE. Among patients, 76 were with MDD, 21 with BD, and 24 with BPD. The pilot study included 23 controls and 14 MDD patients. All participants diagnosed with an MDE were collectively categorized into a single ’Patient’ group for clarity in the following analyses unless otherwise noted.

In the main study, the mean age of the control group was 42 ± 14.07 years, and the mean age of the patient group was 34.7 ± 12.71. 76.66% of the control group were female, compared to 71.07% in the patient group. Regarding employment, 83.3% of the control group were employed full-time, compared to only 9.9% of the patient group. At baseline, the mean PHQ-9 score of the control group was 1.7 ± 1.49. For the patient subcohorts, the baseline PHQ-9 scores were 14.6 ± 5.41 (MDD), 14.57 ± 5.62 (BPD), and 13.53 ± 4.73.

In the pilot study, the mean age of the control group was 30.59 ± 10.54 years, and the mean age of the patient group was 32.38 ± 12.21. 65.21% of the control group were female, compared to 57.14% in the patient group; like the main study, employment, and study engagement were high among the control group, with 95.96% either employed or studying, compared to 42.86% in the patient group. At baseline, the mean PHQ-9 score of the control group was 1.64 ± 1.65, and for the patient group was 13.78 ± 6.09.

#### RQ1: Participation adherence

For this work, we defined adherence as the availability of passively sensed smartphone battery data. The adherence rate over time was measured using survival analysis. Figure 2 shows the survival curve depicting the adherence rate. 65.9% of participants remained in the study through week 8. Adherence rates after week 8 exhibited variability across different groups: 56.7% of the control group, 79% of the BPD group, 69.8% of the MDD group, and 55% of the BD group maintained enrollment. Nonetheless, the log-rank test did not reveal a significant difference in adherence levels between all groups.

**Figure 2:**
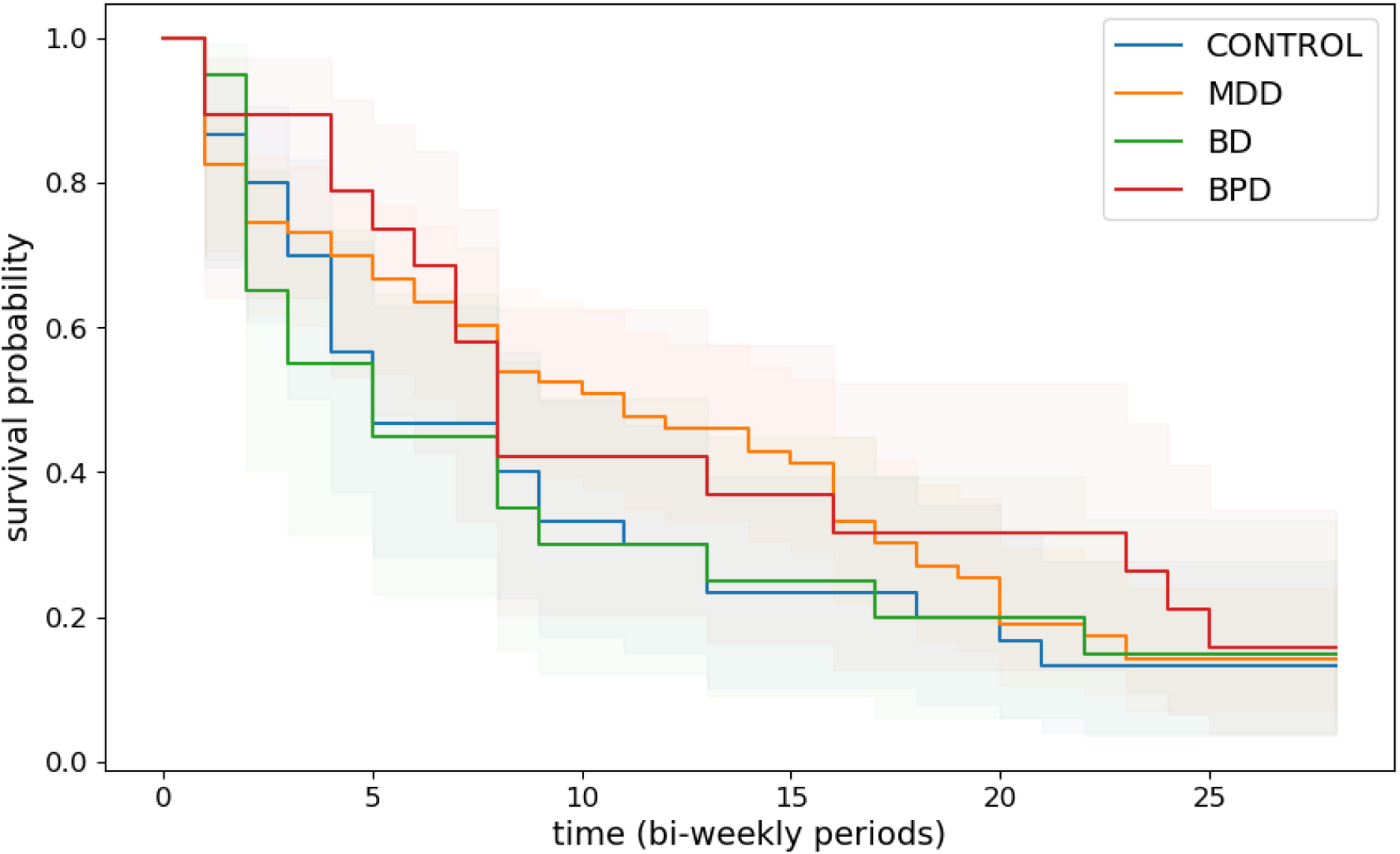
Kaplan-Meier survival curves depicting the probability of continued participation (survival) among different groups in the study. A participant is considered adherent if they provide battery data at least once within each bi-weekly period (14 days), starting from the beginning of the study. Notably, the curves indicate no statistically significant difference in adherence between the groups.

#### RQ2: Quantity and temporal patterns of smartphone usage

To assess the difference between patients and controls in smartphone usage, we measured the quantity of smartphone interaction for each group by analyzing the daily average duration of smartphone screen use and time spent on the top five most-used application categories. To quantify the temporal patterns of smartphone usage, we used the normalized amount of interaction with the screen and these top applications.

The daily average usage of the top five application categories and overall screen time are presented in Table 2. No significant differences were observed in the amount of time spent on each category or the overall volume of smartphone usage between the groups. However, noticeable differences emerged when examining the temporal patterns of application usage. As illustrated in Figure 3, controls tend to use leisure applications more frequently during the weekend, with a peak on Saturday. This pattern suggests that controls prefer to engage with entertainment applications during their free days.

**Figure 3:**
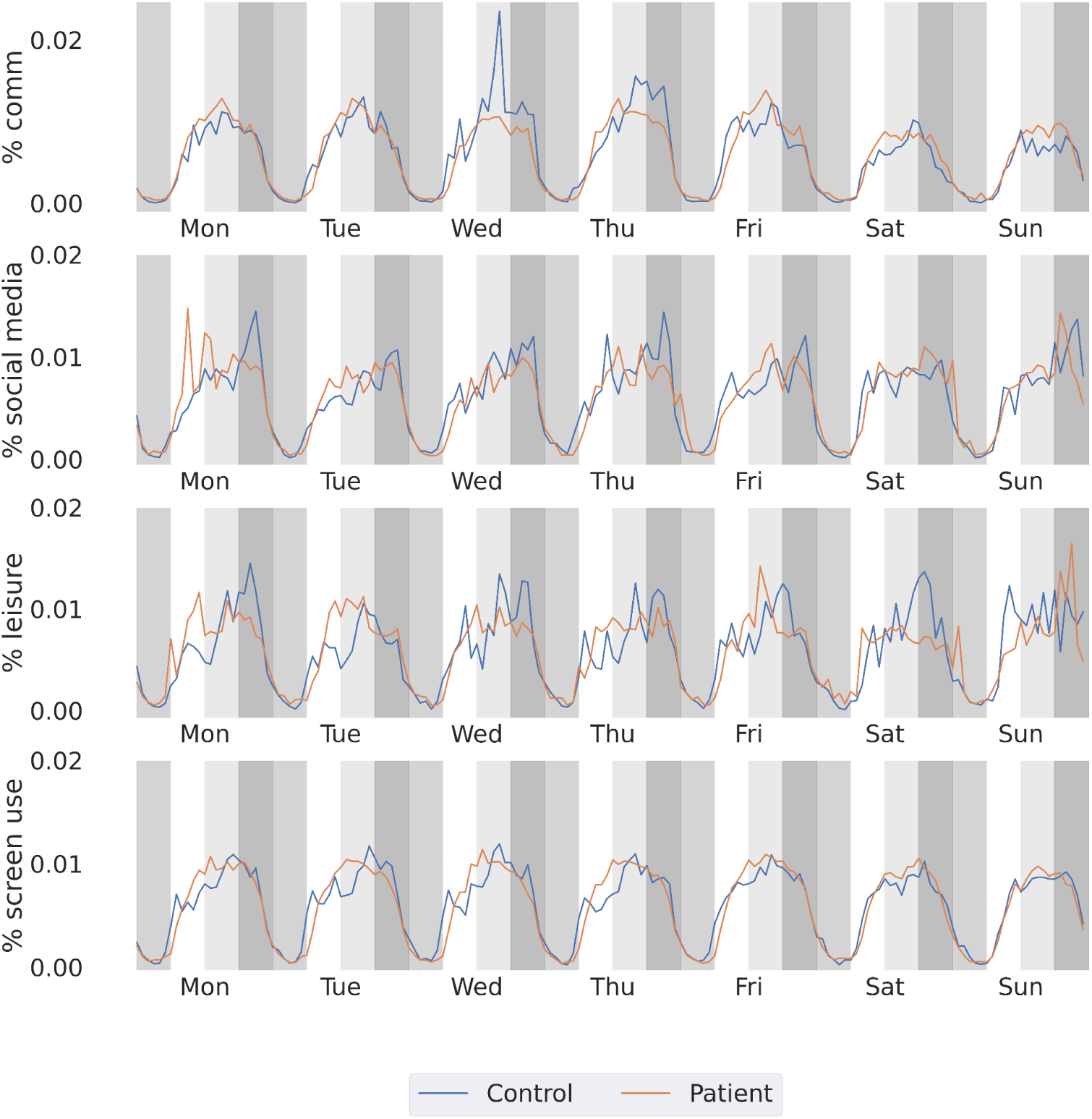
Weekly pattern of application interaction and smartphone usage. Each color region denotes a four-hour daily bin representing night (Midnight—6 AM), morning (6 AM—12 PM), afternoon (12 PM—6 PM), and evening (6 PM—midnight).

**Table 2:** Daily average duration (in seconds) of top 5 app usage categories and total time spent using the smartphone. *P*-values from the MWU test, adjusted to correct for multiple testing.

| Usage duration (seconds) | Control, mean (SD) | Patient , mean (SD) | *P value* |
| --- | --- | --- | --- |
| Communication | 1107.6 (699.0) | 1743.6 (2170.8) | .87 |
| Social media | 1897.2 (1355.4) | 2364.6 (2473.8) | .90 |
| Leisure | 1117.2 (1098.0) | 1752.0 (3055.2) | .87 |
| Games | 690.0 (943.8) | 828.6 (1515.6) | .87 |
| News | 967.2 (2563.8) | 441.6 (919.8) | .90 |
| Screen use | 13554.7<br>(6820.5) | 10690.5<br>(7855) | .55 |

#### RQ3: Between-group difference in the quantity and temporal patterns of behaviors

We employed the same strategy to quantify daily communication and location patterns from smartphone sensors, sleep patterns from bed sensors, activity patterns from actigraphs, and weekly temporal communication patterns.

#### Communication

We used call and SMS data collected by smartphones as a proxy of social activity. We collected 46,788 incoming and outgoing calls, of which 39,491 were longer than zero seconds, with an average call duration of 404.9 seconds. Also, we collected 46,031 text messages within the same period. Table 3 presents the mean (±sd) of the two groups’ daily calls and text messages. A comparison of the daily average communication between groups did not show significant differences.

**Table 3:** Daily average call and text message (SMS) per group for each study. *P*-values from the MWU test were adjusted for multiple tests.

| Features | Main, mean (sd) |  |  | Pilot, mean (sd) |  |  |
| --- | --- | --- | --- | --- | --- | --- |
|  | Control | Patient | *P value* | Control | Patient | *P value* |
| Num. of incoming calls | 0.67<br>(0.58) | 1.26<br>(1.17) | .64 | 0.70<br>(0.34) | 0.80<br>(0.53) | .49 |
| Num. of outgoing calls | 1.26<br>(1.21) | 1.59<br>(1.37) | .90 | 1.32<br>(0.92) | 0.69<br>(0.56) | .15 |
| Incoming call duration (seconds) | 292.2<br>(365.0) | 473.4<br>(441.5) | .64 | 160.5<br>(162.0) | 1134.3<br>(2103.8) | .34 |
| Outgoing call duration (seconds) | 332.8<br>(328.7) | 460.3<br>(596.3) | .94 | 163.1<br>(118.6) | 125.1<br>(165.9) | .49 |
| Num. of incoming SMS | 1.24<br>(0.79) | 1.48<br>(1.23) | .94 | 1.24<br>(1.17) | 0.99<br>(0.48) | 1.0 |
| Num. of outgoing SMS | 0.66<br>(0.98) | 0.95<br>(1.27) | .69 | 0.93<br>(1.24) | 0.91<br>(0.87) | .84 |

Figure 4 presents the weekly communication rhythms for both groups. A visual inspection reveals that both groups tended to have a decreased communication volume during the weekend. Within the control group, call events were more frequently observed in the afternoon bins (12 PM-6 PM).

**Figure 4:**
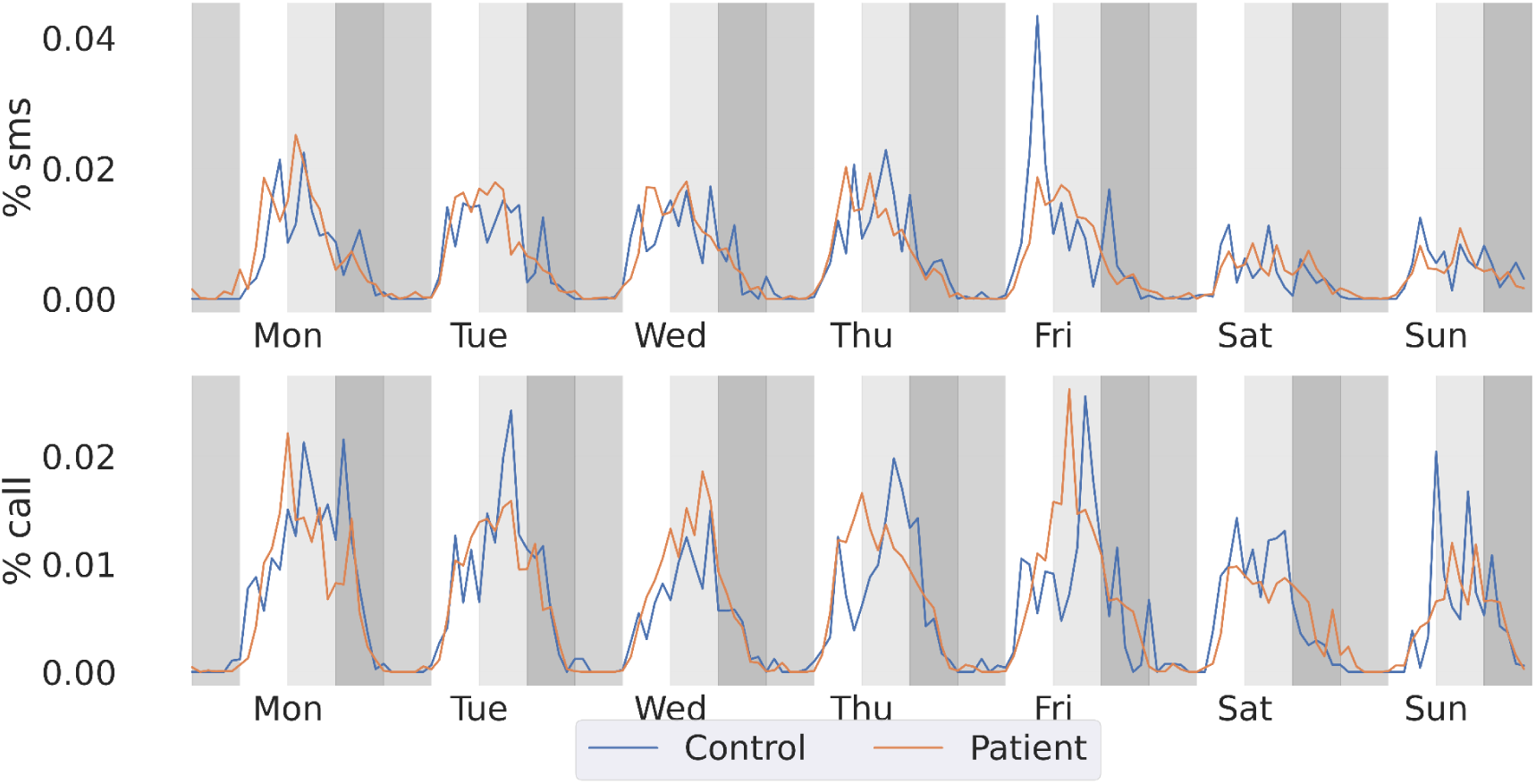
Weekly pattern of communication. Each color region denotes a four-hour daily bin representing night (Midnight - 6 AM), morning (6 AM - 12 PM), afternoon (12 PM - 6 PM), and evening (6 PM-Midnight).

#### Sleep and Physical Activity

We examined all participants’ sleep and physical activity patterns via features extracted from the actigraph and the bed sensor. We look at these two behaviors together as they complement each other throughout the day. We measured the sleep duration from the bed sensor and the active hours from the actigraphy. The daily average active hours and sleep duration of each group in each study are presented in Figure 5. We found no between-group difference in sleep duration in either of the studies (Patient: 7.99 ± 1.80, Control: 8.13 ± 1.05, MWU test *P*-value: .57). Similarly, active hours showed no significant difference between groups (Patient: 13.93 ± 1.57, Control: 14.80 ± 1.30, MWU test *P*-value: .14).

**Figure 5:**
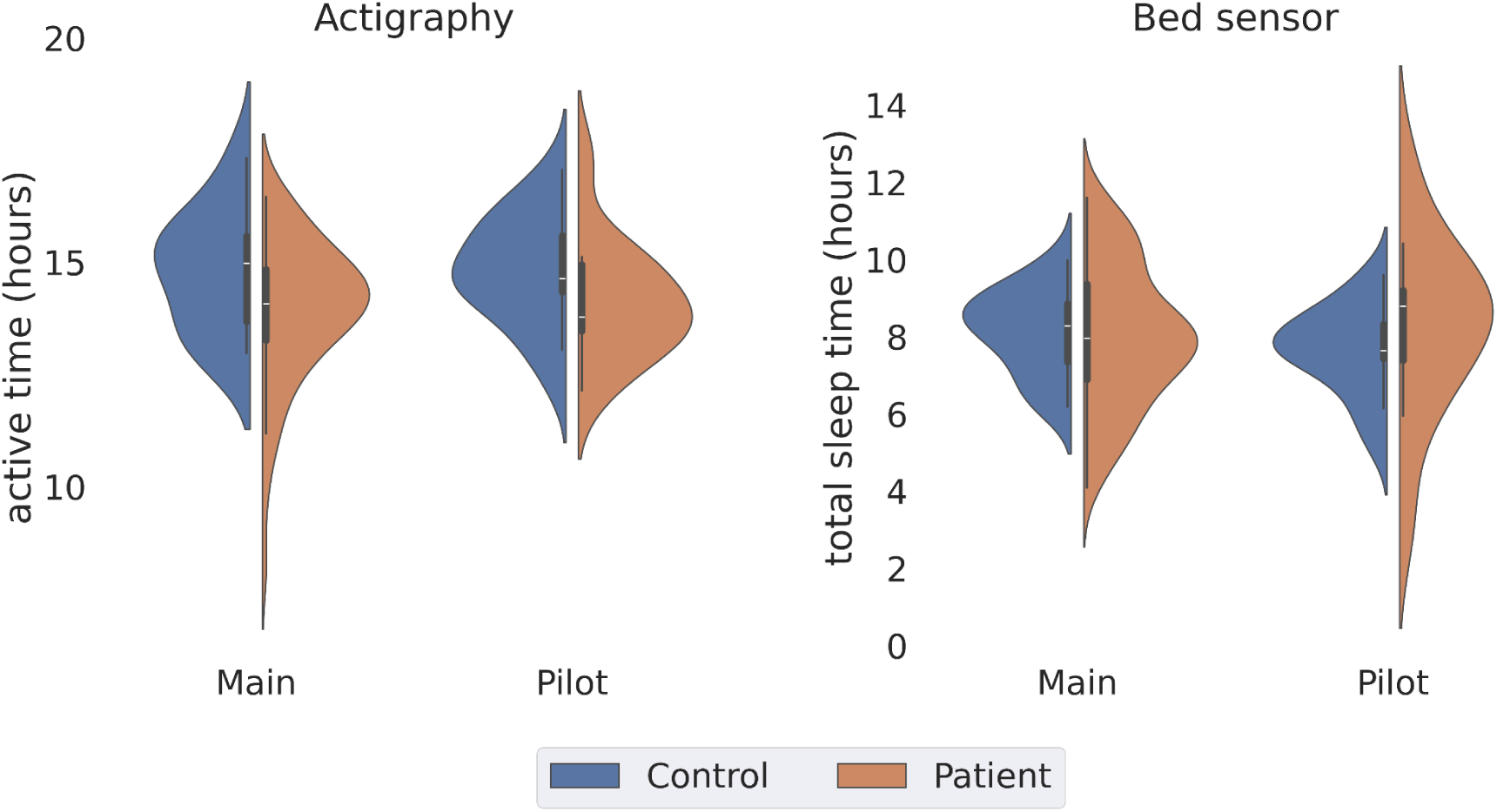
Sleep variables gathered from actigraph and bed sensor.

We measured the intensity of physical activity using the standard deviation of the phone’s magnitude computed from the accelerometer sensor. Again, we found no significant difference in daily physical activity intensity between the groups (Patient: 0.60 ± 0.32, Control: 0.58 ± 0.34, MWU test *p*-value: .35). The temporal trend (Figure 6) showed a general decrease in activity intensity towards the weekend. This reduction is more noticeable in the control group.

**Figure 6.**
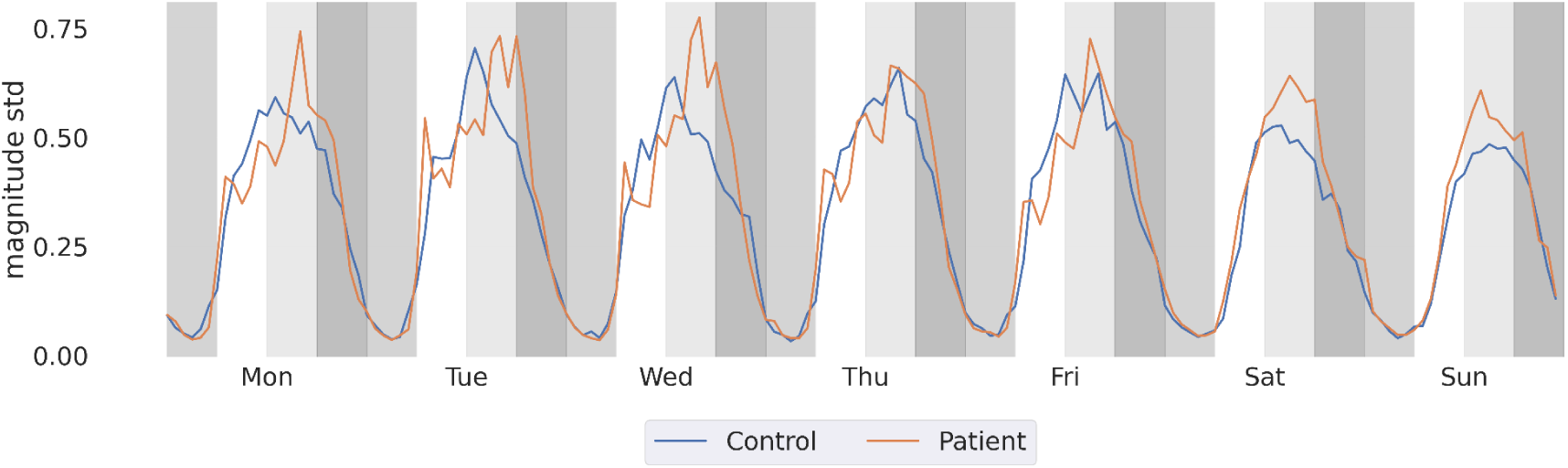
Weekly pattern of physical activity intensity measured by the standard deviation of the accelerometer’s magnitude.

#### Mobility and Location

Due to the inherent difference in mobility patterns during workdays and weekends, we separated the features for these two time frames. Figure 7 demonstrates the weekday location features of the two groups. Significant differences were observed in weekday location features. Specifically, both location variance (Patient: -11.91 ± 2.50, Control: -10.04 ± 2.73, MWU test *P*-value: .004) and normalized entropy (Patient: 1.57 ± 1.10, Control: 2.10 ± 1.38, MWU test *P*-value: .05) were lower in patients. However, no significant differences in weekend location features were detected. When the analysis was restricted to participants with work duties, only the weekday location variance remained significant (Patient: -11.14 ± 2.08, Control: -10.00 ± 2.80, MWU test *P*-value: .05). No significant findings were observed in the Pilot study.

**Figure 7.**
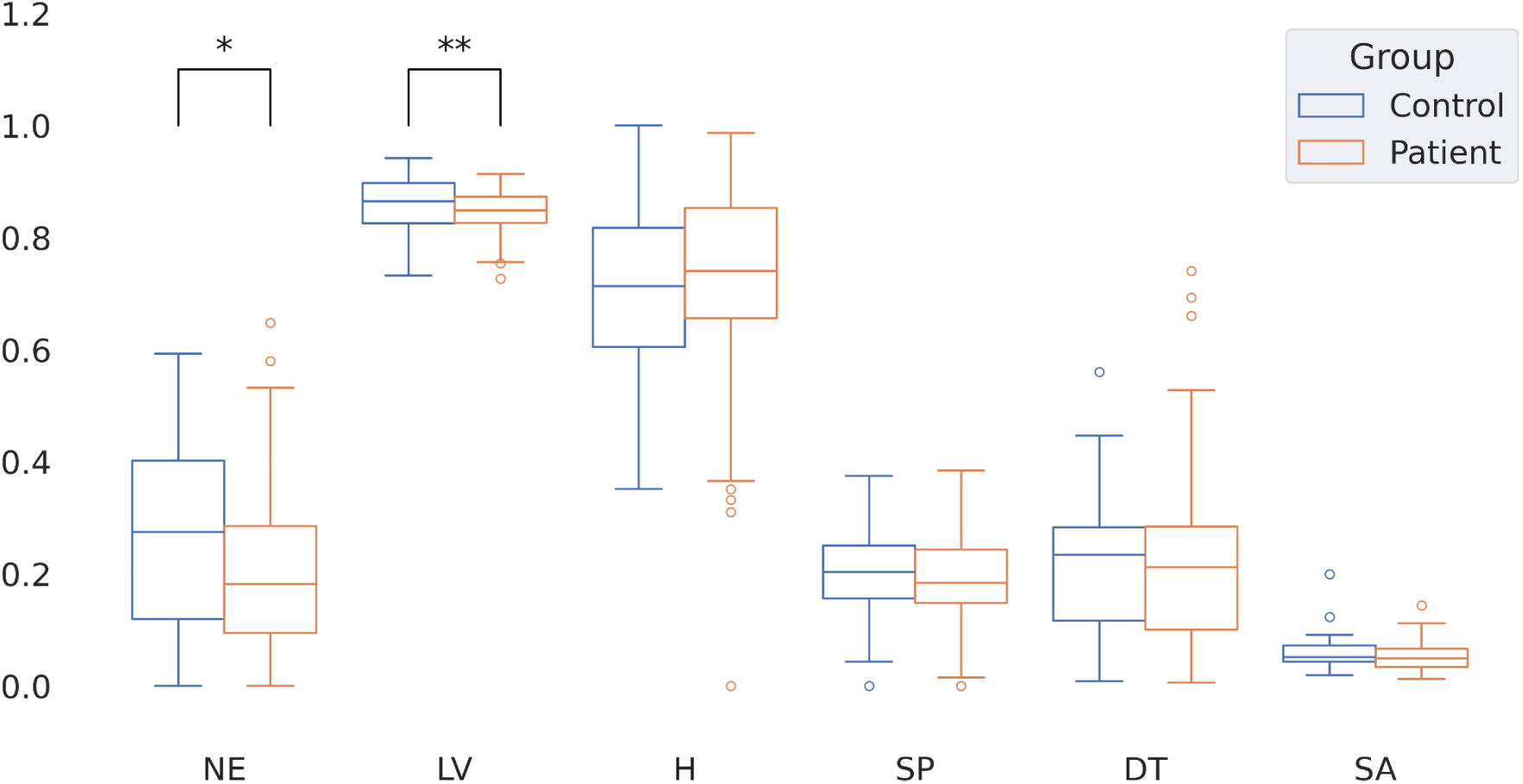
Weekday location features normalized to the [0,1] range. Asterisks denote significance. NE = normalized entropy, LV = location variance, H = proportion of time at home, SP = significant places, DT = distance traveled, SA = speed average.

#### RQ4: Within-group difference in smartphone usage and behavior patterns

We examined the extent to which participants within the same group exhibit different temporal rhythms of smartphone usage patterns. Within-group rhythm similarity measures the consistency of behavioral patterns among individuals within the same group. Higher homogeneity indicates that group members exhibit similar behaviors, reflecting a unified group dynamic. To compare group similarity, we calculate the cosine distance between individuals’ average weekly rhythms within each group (patients-patients and controls-controls). We apply the MWU test to determine significant differences between groups’ mean distances, thus evaluating internal group consistency.

Figure 8 shows the relative within-group distance for each of the groups. Patients established a more homogenous communication rhythm than the controls, meaning that their time allocation for communication was more similar to one another compared to how the healthy controls compare to each other (MWU test, Call: *P*-value: < .001; SMS: *P*-value: .036). Because most patients did not have full-time jobs at the time of the study, we also examined the potential confounders by composing the same analysis only on the participants who had full-time jobs. Nonetheless, the previous results persist, meaning employment did not play a significant role in maintaining communication rhythms.

**Figure 8:**
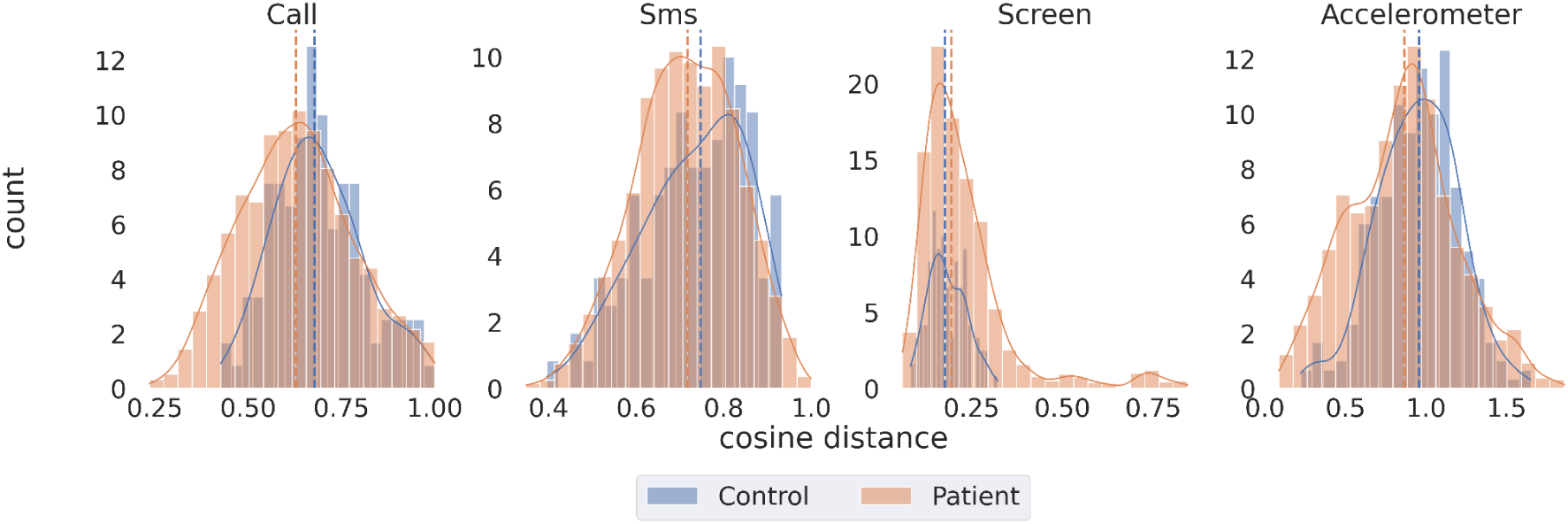
Histogram representing the intra-difference of weekly rhythm measured by cosine distance. Smaller distances indicate more intra-individual similarity.

On the other hand, patients exhibited less consistent smartphone usage rhythms compared to controls (MWU test, P-value: .002). However, among participants with full-time jobs, controls displayed less consistency in smartphone usage than patients (MWU test, P-value: .002). Nevertheless, patients exhibited more consistency in activity rhythms captured by accelerometers than controls, across the entire sample as well as the full-time employed subgroup (MWU test, *P*-value < .001).

#### RQ5: Sensor measures as predictors of presence of depression

Aside from examining the difference in sensor measures between groups of participants, we also measured the association between those measures and the presence of depressive symptoms. There were 398 instances of no or mild symptoms (PHQ-9 score below 10) and 357 instances of moderate to severe symptoms (PHQ-9 score of 10 or higher). Among these, there were 76 occasions when participants transitioned from a mild or no symptoms stage to a severe stage and vice versa. We fitted a GLMM model separately for each sensor and presented the models with significant results in Table 4. All predictors were normalized to the [0,1] range. Due to the prevalence of missing data, location features were imputed using MICE imputation.

**Table 4:** Location and communication factors predicting the severity of depression symptoms. The call durations are in seconds.

|  | Estimates | 95% CI | *P value* |
| --- | --- | --- | --- |
| **Communication** |  |  |  |
| Num. of incoming calls | -0.08 | -5.14 - 4.97 | .97 |
| Num. of outgoing calls | 0.14 | -3.44 - 3.73 | .93 |
| Incoming call duration | 5.33 | -0.08 - 10.75 | .053* |
| Outgoing call duration | -7.93 | -13.17 - -2.69 | .003** |
| Num. of incoming SMS | 0.27 | -5.37 - 5.92 | .92 |
| Num. of outgoing SMS | 2.74 | -2.58 - 8.06 | .31 |
| **Location** |  |  |  |
| Location variance | -7.5 | -17.06 - 1.87 | .11 |
| Normalized entropy | 0.20 | -3.24 - 3.64 | .90 |
| Distance travelled | 1.75 | -2.18 - 5.70 | .38 |
| SPs | 1.35 | -1.00 - 1.60 | .65 |
| Time at home | 5.16 | 0.78 - 9.54 | .02* |
| Speed average | 0.05 | -1.03 - 1.14 | .92 |

Our analysis of sensor data revealed notable associations in communication and location patterns related to depression symptoms. For communication patterns, the duration of incoming calls was marginally positively associated with depression symptoms (beta: 5.33, 95% CI: -0.08 to 10.75, *P*-value: .053). In contrast, the duration of outgoing calls showed a negative correlation (beta: -7.93, 95% CI: -13.17 to -2.69, *P*-value: .003), indicating that longer outgoing calls were linked to the presence of depression. Among all location features, the proportion of time spent at home was positively associated with depression presence (beta: 5.16, 95% CI: 0.78 to 9.54, *P*-value: .02), suggesting a bi-directional relationship between increased time at home and the presence of depression symptoms. Detailed results for the other measures are presented in Appendix 2.

## Discussion

### Principal Results

This paper introduced the MoMo-Mood Pilot and MoMo-Mood studies, which comprise a rich, longitudinal multisensor behavioral dataset collected from multiple cohorts. It presented the study setting, its deployment, and data characteristics.

In the MoMo-Mood study, there were no significant differences in adherence between patients and healthy controls. Our analysis of smartphone usage patterns did not reveal significant differences between the groups’ daily quantity of smartphone usage. However, the weekly temporal patterns revealed distinct preferences, such that the control group typically made or received calls in the afternoon. Furthermore, we initially hypothesized that patients would exhibit notable differences in activities and behaviors crucial for assessing mood disorder symptoms or associated psychosocial disability across four domains: social activity, sleep, physical activity, and mobility. Our findings partially supported this hypothesis, as we found significant differences in mobility and location patterns, particularly in terms of normalized entropy and variance of locations during weekdays. However, the groups showed no notable differences in sleep, social, and physical activity patterns.

The intra-group temporal rhythm analysis revealed more nuanced findings. The patient group demonstrated more consistent communication and physical activity rhythms than the control group. On the other hand, patients showed more varied rhythms in smartphone screen use. Interestingly, this trend for the smartphone screen patterns, reversed when we took employment status into account.

Using passive data to predict the presence of depressive symptoms; we observed a positive association between the duration of incoming calls and the proportion of time spent at home. Conversely, the duration of outgoing calls was negatively associated with the presence of depressive symptoms.

### Implications

The behavioral data reveals that control and patient groups actively use their smartphones for similar amounts of time. This similarity in smartphone engagement validates the potential of smartphone data as a reliable source for finding new behavioral markers. Noteworthy, even while the patients were mildly to moderately clinically depressed, they were actively engaged with their smartphones. Further, the analysis supports the utility of smartphone data in capturing behavioral patterns, showing disparities in behavioral rhythms, including communication and screen usage patterns.

Furthermore, our study did not confirm the significance of sleep duration in detecting depression symptoms, in contrast to prior research [10,49,50]. While the study did not find a difference in sleep duration between depressed patients and the healthy controls, it is well-known that both insomnia and hypersomnia are symptoms of depression [21], which we did not measure in this study. In the future, we will conduct a more fine-grained analysis focusing on the disturbances and individual differences in sleep.

Our exploration of weekday and weekend mobility patterns and location features revealed a significant difference in weekday normalized entropy and location variance between the groups. These findings are consistent with prior research, such as the study by Saeb et al. [23,24], which identified a significant correlation between depression scores and location features, including location variance and location entropy. The importance of mobility and location data is also supported by previous literature [10,51,52], recognizing these data as crucial components in modeling depression severity.

Investigating the similarity of the behavior of patients to one another and the healthy controls to one another, we saw that across three different activity types, communication via calls and SMS, and smartphone acceleration, patients behave more similarly to one another. These findings indicate that the rhythms of different activities and how patients allocate their time to these activities throughout the week are associated with their mental health condition. These results persisted even when controlling for the subject’s work status. This highlights the importance of temporality and temporal behavioral features as behavioral markers for mood disorders.

It is essential to keep in mind that digital phenotyping studies aim to capture subtle differences and patterns in the behavior of patients and healthy controls, which can possibly be missed even by trained clinicians. Any major behavioral differences between these groups have naturally already been noticed and studied by experts in psychiatry. Our study adds to the body of knowledge in digital phenotyping. It points out essential yet subtle features that need further research and can potentially be used to expand the behavioral symptomology of mood disorders.

Our findings provide valuable longitudinal insights and generalizability by representing different types of major depressive patients. It is important to note that most of our patients suffered from mild to moderate clinical depression, representing the range of severity of the majority of outpatients with depression in health care. However, the inclusion of psychiatric inpatients with severe or psychotic depressive patients or bipolar patients with manic episodes might have revealed more marked deviations from standard behavioral patterns. Furthermore, patients were receiving treatment, and it is likely that, for example, the observed sleep symptoms were influenced by ongoing pharmacotherapy.

### Limitations

The patient cohorts included 147 major depressive patients and 54 healthy controls. While our sample is comparable to or even larger than those in similar studies, it remains moderate. This limitation, however, is offset by the extended duration of data collection.

Further, the data is limited by the decreasing participant adherence and increasing drop-out rate as the study progressed. Technical issues, the burden of participation, lack of motivation, or health status may cause poor adherence. Other digital phenotyping studies have similarly reported adherence of various degrees, from 65.3% smartphone data completeness (study with 334 MDD patients covering 12 weeks [50]) to 99% completeness (29 patients diagnosed with BP monitored for one year [53]). The adherence may also vary between the study cohorts. For example, a study [52] reported average adherence levels of 66,8% for patients with BD and 84,8% for the healthy controls for daily smartphone-reported self-assessments collected over nine months. However, in our study, adherence remained similar for different subcohorts. In our study, participants were not followed up on or engaged with after they completed the active phase of the study (approximately two weeks after the enrollment). They received four movie tickets for participation in the study, two at the time of enrolment and two after completing the active phase. They did not receive any further compensation or feedback. The lack of further follow-up or feedback to participants has likely played an essential role in our dropout rates.

Our study is limited by the data types that we can collect. It thus does not capture the full extent of a person’s communication (e.g., communication via social media or messaging applications). Furthermore, for devices like bed sensors and actigraphs, we rely on the pre-processed data provided by the manufacturer, which can potentially introduce unknown errors and biases.

### Future research

We have collected a very rich dataset that requires much more extensive exploration. This paper provides an overview of the study and compares behavioral patterns at the group level for the passively collected data from smartphones, bed sensors, and actigraphs. Future research will need to cover more fine-grained analysis at the individual level, a more profound investigation of parameters deemed important in this work, and the link between passive data and subjective data collected through validated clinical questionnaires.

The results of this study show that the mobile behavioral data, including features from smartphone usage, communication, location, mobility, and sleep, collected within this study can be further used to predict and monitor the patients’ depression. The revealed group-level differences observed in this study by comparing the rhythms suggest focusing on extracting temporal features from the behavioral data measuring the rhythms and temporal patterns. Thus, further studies are required to validate these findings. We have released an open-source behavioral data analysis toolbox, Niimpy [45], to facilitate the studies and encourage researchers to utilize and further develop it. Furthermore, we propose that future analyses utilize personalized statistical and machine learning models, accounting for the differences in behavior and sociodemographic information (for example, subjects’ age, sex, and work status). In the future, we will extend our analyses and explore the possibility of predicting the future mood state and severity of depression from features extracted from the passively collected data.

## Conclusions

This study demonstrates the feasibility of harnessing data from a cohort of patients with different types of clinically diagnosed depressive syndromes. It also shows the important features and data streams for future analyses of behavioral markers of mood disorders. However, among outpatients with mild to moderate depressive disorders investigated, their group-level differences from healthy controls in any of the modalities alone remain overall modest, even if within-individual variations in behavioral patterns related to the presence of depression could be demonstrated. Overall, the sensors used in passive monitoring may more readily detect gross observable behavioral abnormalities, which may emerge only in the epidemiologically more rare severe range of depression or in mania. Therefore, future studies need to be able to combine data from multiple domains and modalities to detect more subtle differences, identify individualized signatures, and combine passive data with clinical ratings.

## Data Availability

The data is not publicly available.

## Acknowledgments

We acknowledge the computational resources provided by the Aalto Science-IT project. TA acknowledges the support of the James S. McDonnell Foundation. RD and TA acknowledge helpful discussions with Denzil Ferreira, Niels van Berkel, and others in the Center for Ubiquitous Computing, University of Oulu.

## Conflicts of Interest

The authors declare no conflicts of interest.

## Abbreviations

MoMo-Mood: Mobile Monitoring of Mood
MDE: Major depressive episode
MDD: Major depressive disorder
BD: Bipolar disorder
BPD: Borderline personality disorder
App: Phone application
sd: Standard deviation
MWU test: Mann-Whitney U test
NE: Normalized entropy
LV: Location variance
H: Proportion of time at home
SP: Significant places
DT: Distance traveled
SA: Speed average

## Appendix 1

A significant challenge in collecting GPS data is the prevalence of missing data and outlier values. Less than 1% of the data points consisted of the *(longitude, latitude) = (0, 0)* coordinate, which fell in the middle of the Atlantic Ocean and was not a plausible location for our participants. Furthermore, a proportion of data points derived from the Wi-Fi signal contain very high-speed values (>10^2^ m/s). Therefore, we retained only data points derived from GPS sources and systematically eliminated outliers to ensure data validity. Following this process, 6,287,582 data points were filtered out from the original count of 11,346,218, resulting in a total of 5,058,636 data points being retained and eligible for further analysis. For the distance traveled feature, we filter the outliers by accepting only daily trips less than 100km.

**Table A1:**
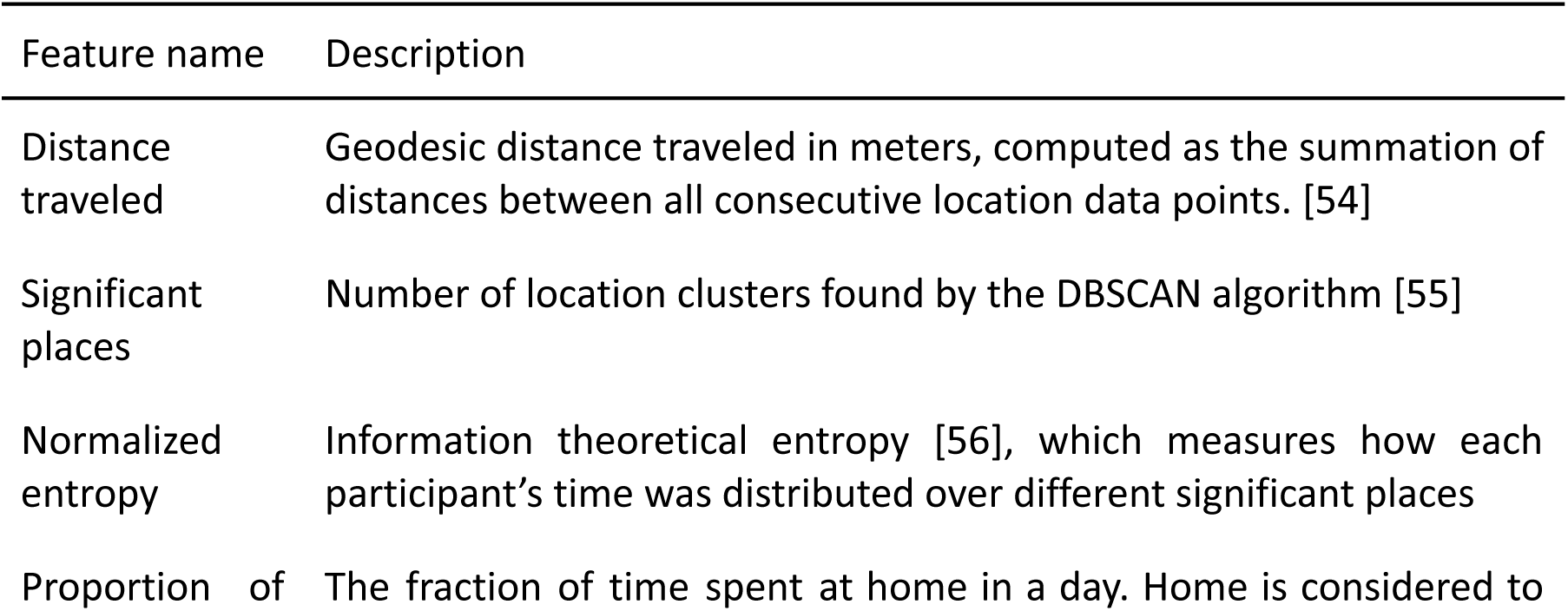
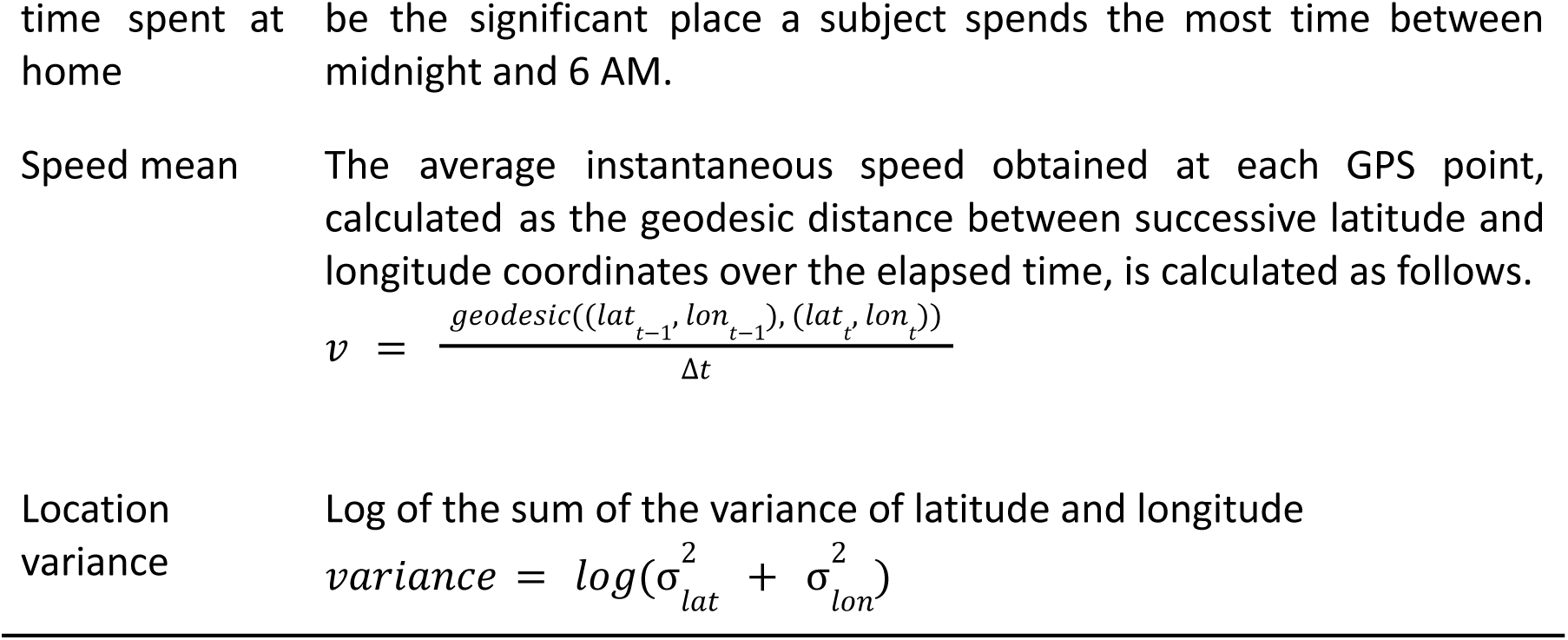
Location and mobility features description.

## Appendix 2

**Table A2:** Description of location features. Weekday and weekend features were calculated separately for each group. The *p*-values are from MWU tests.

|  | Weekday |  |  | Weekend |  |  |
| --- | --- | --- | --- | --- | --- | --- |
|  | Control | Patient | *P value* | Control | Patient | *P value* |
| location variance | -10.22 ± 2.70 | -11.97 ± 2.56 | .006** | -13.08 ± 2.83 | -12.49 ± 3.23 | .34 |
| normalized entropy | 2.05 ± 1.39 | 1.52 ± 1.12 | .05* | 1.07 ± 0.89 | 1.35 ± 1.14 | .34 |
| % time at home | 0.71 ± 0.18 | 0.73 ± 0.18 | .57 | 0.83 ± 0.12 | 0.76 ± 0.18 | .15 |
| number of SPs | 1.62 ± 0.65 | 1.49 ± 0.65 | .30 | 1.31 ± 0.42 | 1.41 ± 0.63 | .40 |
| average speed (km/h) | 14.32 ± 8.35 | 12.48 ± 5.91 | .36 | 12.66 ± 8.58 | 10.88 ± 6.00 | .34 |
| total distance (km) | 11.69 ± 7.29 | 11.83 ± 7.97 | .91 | 8.32 ± 5.99 | 11.32 ± 9.75 | .19 |

## Appendix 3

**Table A3:**
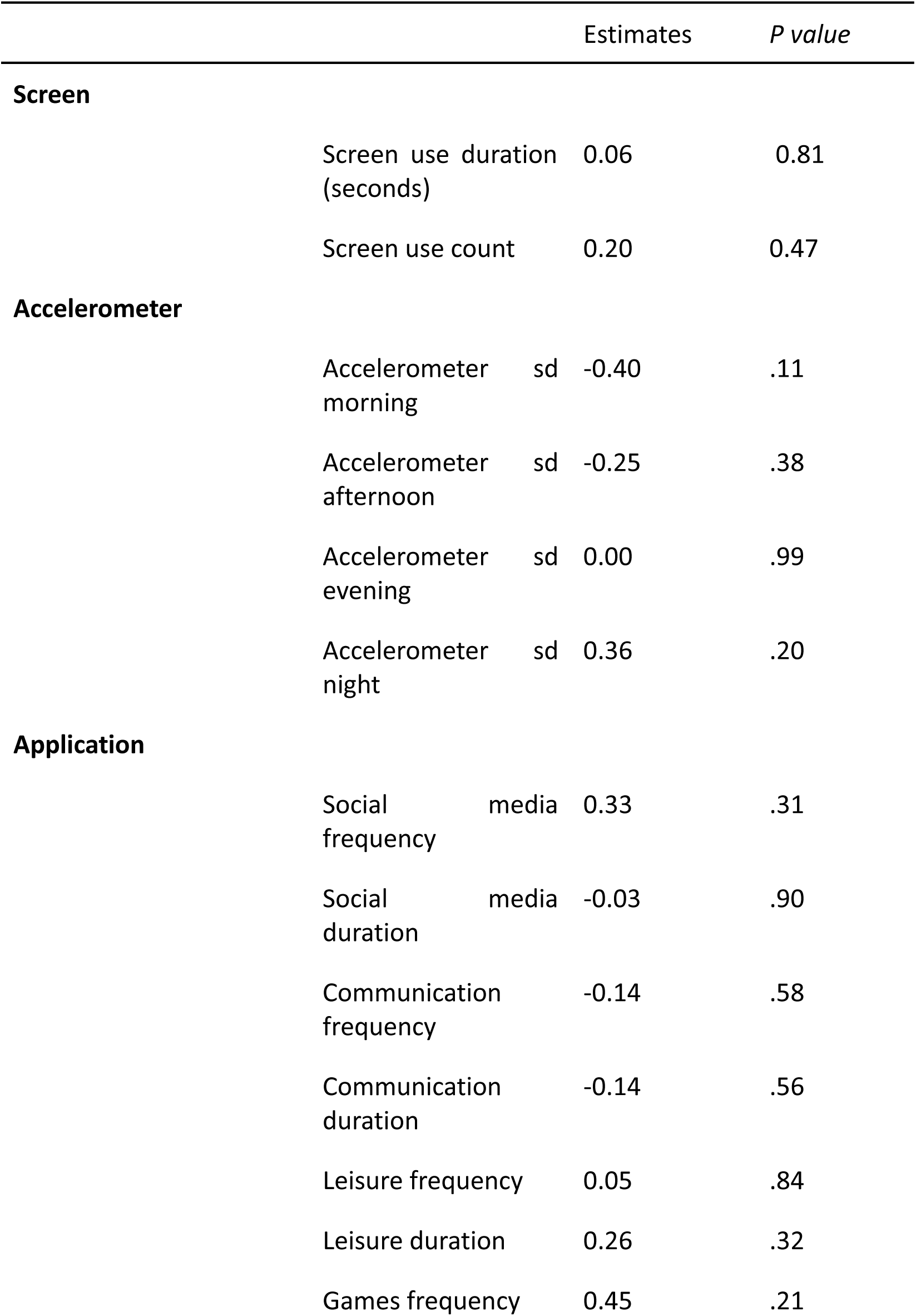
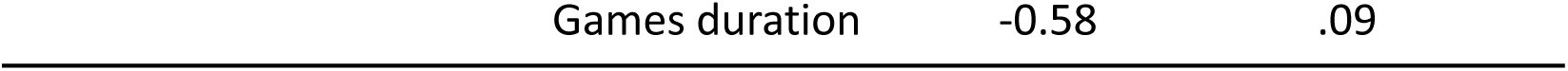
Results of screen usage, accelerometer measurements, and application usage predicting the presence of depressive symptoms.

## Notes

### Competing Interest Statement

The authors have declared no competing interest.

### Funding Statement

This study did not receive any funding.

### Author Declarations

The research protocol for both MoMo-Mood Pilot and MoMo-Mood was approved by the Helsinki and Uusimaa Hospital District (HUS) Ethics Committee and was granted research permits by HUS Psychiatry.

## References

1. Vos T, Lim SS, Abbafati C, et al. Global burden of 369 diseases and injuries in 204 countries and territories, 1990–2019: a systematic analysis for the Global Burden of Disease Study 2019. The Lancet. 2020;396(10258):1204-1222. doi:10.1016/S0140-6736(20)30925-9

2. GBD 2019 Mental Disorders Collaborators. Global, regional, and national burden of 12 mental disorders in 204 countries and territories, 1990–2019: a systematic analysis for the Global Burden of Disease Study 2019. Lancet Psychiatry. 2022;9(2):137-150. doi:10.1016/S2215-0366(21)00395-3

3. Walker ER, McGee RE, Druss BG. Mortality in mental disorders and global disease burden implications: a systematic review and meta-analysis. JAMA Psychiatry. 2015;72(4):334–341. doi:10.1001/jamapsychiatry.2014.2502

4. Plana-Ripoll O, Musliner KL, Dalsgaard S, et al. Nature and prevalence of combinations of mental disorders and their association with excess mortality in a population-based cohort study. World Psychiatry Off J World Psychiatr Assoc WPA. 2020;19(3):339–349. doi:10.1002/wps.20802

5. Costello EJ, Pine DS, Hammen C, et al. Development and natural history of mood disorders. Biol Psychiatry. 2002;52(6):529–542. doi:10.1016/S0006-3223(02)01372-0

6. Ebner-Priemer UW, Trull TJ. Ecological momentary assessment of mood disorders and mood dysregulation. Psychol Assess. 2009;21(4):463–475. doi:10.1037/a0017075

7. Solhan M, Trull T, Jahng S, Wood P. Clinical Assessment of Affective Instability: Comparing EMA Indices, Questionnaire Reports, and Retrospective Recall. Psychol Assess. 2009;21:425–436. doi:10.1037/a0016869

8. Majumder S, Deen MJ. Smartphone Sensors for Health Monitoring and Diagnosis. Sensors. 2019;19(9):2164. doi:10.3390/s19092164

9. Xu X, Chikersal P, Doryab A, et al. Leveraging Routine Behavior and Contextually-Filtered Features for Depression Detection among College Students. Proc ACM Interact Mob Wearable Ubiquitous Technol. 2019;3(3):1–33. doi:10.1145/3351274

10. Xu X, Chikersal P, Dutcher JM, et al. Leveraging Collaborative-Filtering for Personalized Behavior Modeling: A Case Study of Depression Detection among College Students. Proc ACM Interact Mob Wearable Ubiquitous Technol. 2021;5(1):1–27. doi:10.1145/3448107

11. Faurholt-Jepsen M, Bauer M, Kessing LV. Smartphone-based objective monitoring in bipolar disorder: status and considerations. Int J Bipolar Disord. 2018;6(1):6. doi:10.1186/s40345-017-0110-8

12. Barnett I, Torous J, Staples P, Sandoval L, Keshavan M, Onnela JP. Relapse prediction in schizophrenia through digital phenotyping: a pilot study. Neuropsychopharmacology. 2018;43(8):1660–1666. doi:10.1038/s41386-018-0030-z

13. Busk J, Faurholt-Jepsen M, Frost M, Bardram JE, Kessing LV, Winther O. Forecasting Mood in Bipolar Disorder From Smartphone Self-assessments: Hierarchical Bayesian Approach. JMIR MHealth UHealth. 2020;8(4):e15028. doi:10.2196/15028

14. Ikäheimonen A, Luong N, Baryshnikov I, et al. Predicting and Monitoring Symptoms in Diagnosed Depression Using Mobile Phone Data: An Observational Study. Published online June 17, 2024:2024.06.15.24308981. doi:10.1101/2024.06.15.24308981

15. Kroenke K, Spitzer RL, Williams JBW. The PHQ-9. J Gen Intern Med. 2001;16(9):606–613. doi:10.1046/j.1525-1497.2001.016009606.x

16. Maatoug R, Oudin A, Adrien V, et al. Digital phenotype of mood disorders: A conceptual and critical review. Front Psychiatry. 2022;13:895860. doi:10.3389/fpsyt.2022.895860

17. Onnela JP. Opportunities and challenges in the collection and analysis of digital phenotyping data. Neuropsychopharmacology. 2021;46(1):45–54. doi:10.1038/s41386-020-0771-3

18. Torous J, Chan SR, Yee-Marie Tan S, et al. Patient Smartphone Ownership and Interest in Mobile Apps to Monitor Symptoms of Mental Health Conditions: A Survey in Four Geographically Distinct Psychiatric Clinics. JMIR Ment Health. 2014;1(1):e5. doi:10.2196/mental.4004

19. Young AS, Cohen AN, Niv N, et al. Mobile Phone and Smartphone Use by People with Serious Mental Illness. Psychiatr Serv Wash DC. 2020;71(3):280–283. doi:10.1176/appi.ps.201900203

20. Ge L, Yap CW, Ong R, Heng BH. Social isolation, loneliness and their relationships with depressive symptoms: A population-based study. PLOS ONE. 2017;12(8):e0182145. doi:10.1371/journal.pone.0182145

21. American Psychiatric Association, American Psychiatric Association, eds. Diagnostic and Statistical Manual of Mental Disorders: DSM-5. 5th ed. American Psychiatric Association; 2013.

22. Targum SD, Fava M. Fatigue as a Residual Symptom of Depression. Innov Clin Neurosci. 2011;8(10):40–43.

23. Saeb S, Lattie EG, Schueller SM, Kording KP, Mohr DC. The relationship between mobile phone location sensor data and depressive symptom severity. PeerJ. 2016;4:e2537. doi:10.7717/peerj.2537

24. Saeb S, Zhang M, Karr CJ, et al. Mobile Phone Sensor Correlates of Depressive Symptom Severity in Daily-Life Behavior: An Exploratory Study. J Med Internet Res. 2015;17(7):e4273. doi:10.2196/jmir.4273

25. Chow PI, Fua K, Huang Y, et al. Using Mobile Sensing to Test Clinical Models of Depression, Social Anxiety, State Affect, and Social Isolation Among College Students. J Med Internet Res. 2017;19(3):e6820. doi:10.2196/jmir.6820

26. Nutt D, Wilson S, Paterson L. Sleep disorders as core symptoms of depression. Dialogues Clin Neurosci. 2008;10(3):329–336. doi:10.31887/DCNS.2008.10.3/dnutt

27. Aledavood T, López E, Roberts SGB, et al. Daily Rhythms in Mobile Telephone Communication. PLOS ONE. 2015;10(9):e0138098. doi:10.1371/journal.pone.0138098

28. Aledavood T, Lehmann S, Saramäki J. Digital daily cycles of individuals. Front Phys. 2015;3. Accessed June 2, 2022. https://www.frontiersin.org/article/10.3389/fphy.2015.00073

29. Aledavood T, Kivimäki I, Lehmann S, Saramäki J. Quantifying daily rhythms with non-negative matrix factorization applied to mobile phone data. Sci Rep. 2022;12(1):5544. doi:10.1038/s41598-022-09273-y

30. Aledavood T, López E, Roberts SGB, et al. Channel-Specific Daily Patterns in Mobile Phone Communication. In: Battiston S, De Pellegrini F, Caldarelli G, Merelli E, eds. *Proceedings of ECCS* 2014. Springer Proceedings in Complexity. Springer International Publishing; 2016:209-218. doi:10.1007/978-3-319-29228-1_18

31. Luong N, Barnett I, Aledavood T. The impact of the COVID-19 pandemic on daily rhythms. J Am Med Inform Assoc. 2023;30(12):1943–1953. doi:10.1093/jamia/ocad140

32. Aledavood T, Lehmann S, Saramäki J. Social network differences of chronotypes identified from mobile phone data. EPJ Data Sci. 2018;7(1):46. doi:10.1140/epjds/s13688-018-0174-4

33. Braund TA, Zin MT, Boonstra TW, et al. Smartphone Sensor Data for Identifying and Monitoring Symptoms of Mood Disorders: A Longitudinal Observational Study. JMIR Ment Health. 2022;9(5):e35549. doi:10.2196/35549

34. Aledavood T, Torous J, Triana Hoyos AM, Naslund JA, Onnela JP, Keshavan M. Smartphone-Based Tracking of Sleep in Depression, Anxiety, and Psychotic Disorders. Curr Psychiatry Rep. 2019;21(7):49. doi:10.1007/s11920-019-1043-y

35. Sheehan DV, Lecrubier Y, Sheehan KH, et al. The Mini-International Neuropsychiatric Interview (M.I.N.I.): the development and validation of a structured diagnostic psychiatric interview for DSM-IV and ICD-10. J Clin Psychiatry. 1998;59 Suppl 20:22-33;quiz 34-57.

36. First MB, Gibbon M. The Structured Clinical Interview for DSM-IV Axis I Disorders (SCID-I) and the Structured Clinical Interview for DSM-IV Axis II Disorders (SCID-II). In: *Comprehensive Handbook of Psychological Assessment, Vol. 2: Personality Assessment*. John Wiley & Sons, Inc.; 2004:134-143.

37. Stone AA, Shiffman SS, DeVries MW. Ecological momentary assessment. In: *Well-Being: The Foundations of Hedonic Psychology*. Russell Sage Foundation; 1999:26-39.

38. Aledavood T, Hoyos AMT, Alakörkkö T, et al. Data Collection for Mental Health Studies Through Digital Platforms: Requirements and Design of a Prototype. JMIR Res Protoc. 2017;6(6):e6919. doi:10.2196/resprot.6919

39. Ferreira D, Kostakos V, Dey AK. AWARE: Mobile Context Instrumentation Framework. Front ICT. 2015;2. Accessed February 16, 2024. https://www.frontiersin.org/articles/10.3389/fict.2015.00006

40. Baryshnikov I, Aledavood T, Rosenström T, et al. Relationship between daily rated depression symptom severity and the retrospective self-report on PHQ-9: A prospective ecological momentary assessment study on 80 psychiatric outpatients. J Affect Disord. 2023;324:170–174. doi:10.1016/j.jad.2022.12.127

41. Servia-Rodríguez S, Rachuri KK, Mascolo C, Rentfrow PJ, Lathia N, Sandstrom GM. Mobile Sensing at the Service of Mental Well-being: a Large-scale Longitudinal Study. In: Proceedings of the 26th International Conference on World Wide Web. International World Wide Web Conferences Steering Committee; 2017:103-112. doi:10.1145/3038912.3052618

42. Ravi N. Activity Recognition from Accelerometer Data. Aaai. 2005;5:1541–1546.

43. Oakley NR. Validation with polysomnography of the Sleepwatch sleep/wake scoring algorithm used by the Actiwatch activity monitoring system. Mini Mitter Co. Sleep. 2:0–140.

44. Roenneberg T, Allebrandt KV, Merrow M, Vetter C. Social Jetlag and Obesity. Curr Biol. 2012;22(10):939-943. doi:10.1016/j.cub.2012.03.038

45. Ikäheimonen A, Triana AM, Luong N, et al. Niimpy: A toolbox for behavioral data analysis. SoftwareX. 2023;23. doi:10.1016/j.softx.2023.101472

46. Kaplan EL, Meier P. Nonparametric Estimation from Incomplete Observations. J Am Stat Assoc. 1958;53(282):457–481. doi:10.1080/01621459.1958.10501452

47. Raudenbush SW, Bryk AS. Hierarchical Linear Models: Applications and Data Analysis Methods. SAGE; 2002.

48. Benjamini Y, Hochberg Y. Controlling the False Discovery Rate: A Practical and Powerful Approach to Multiple Testing. J R Stat Soc Ser B Methodol. 1995;57(1):289–300. doi:10.1111/j.2517-6161.1995.tb02031.x

49. Chikersal P, Doryab A, Tumminia M, et al. Detecting Depression and Predicting its Onset Using Longitudinal Symptoms Captured by Passive Sensing: A Machine Learning Approach With Robust Feature Selection. ACM Trans Comput-Hum Interact. 2021;28(1):3:1-3:41. doi:10.1145/3422821

50. Bai R, Xiao L, Guo Y, et al. Tracking and Monitoring Mood Stability of Patients With Major Depressive Disorder by Machine Learning Models Using Passive Digital Data: Prospective Naturalistic Multicenter Study. JMIR MHealth UHealth. 2021;9(3):e24365. doi:10.2196/24365

51. Laiou P, Kaliukhovich DA, Folarin AA, et al. The Association Between Home Stay and Symptom Severity in Major Depressive Disorder: Preliminary Findings From a Multicenter Observational Study Using Geolocation Data From Smartphones. JMIR MHealth UHealth. 2022;10(1):e28095. doi:10.2196/28095

52. Faurholt-Jepsen M, Busk J, Vinberg M, et al. Daily mobility patterns in patients with bipolar disorder and healthy individuals. J Affect Disord. 2021;278:413–422. doi:10.1016/j.jad.2020.09.087

53. Ebner-Priemer UW, Mühlbauer E, Neubauer AB, et al. Digital phenotyping: towards replicable findings with comprehensive assessments and integrative models in bipolar disorders. Int J Bipolar Disord. 2020;8(1):35. doi:10.1186/s40345-020-00210-4

54. Karney CFF. Algorithms for geodesics. J Geod. 2013;87(1):43-55. doi:10.1007/s00190-012-0578-z

56. Shannon CE, Weaver W. The Mathematical Theory of Communication. University of Illinois Press; 1949.

